# Real-time clinician text feeds from electronic health records

**DOI:** 10.1101/2020.10.02.20205617

**Authors:** James Teo, Vlad Dinu, William Bernal, Phil Davidson, Vitaliy Oliynyk, Cormac Breen, Richard D Barker, Richard Dobson

## Abstract

Analyses of search engine and social media feeds have been attempted for infectious disease outbreaks^1^, but have been found to be susceptible to artefactual distortions from health scares or keyword spamming in social media or the public internet ^2–4^. We describe an approach using real-time aggregation of keywords and phrases of free text from real-time clinician-generated documentation in electronic health records to produce a customisable real-time viral pneumonia signal providing up to 2 days warning for secondary care capacity planning. This low-cost approach is open-source, is locally customisable, is not dependent on any specific electronic health record system and can be deployed at multiple organisational scales.

## Introduction

Analyses of search engine and social media feeds have been attempted for infectious disease outbreaks ^1^, but have been susceptible to artefactual distortions from health scares or keyword spamming in social media or the public internet ^2–4^. Electronic health records have a built-in control for these distortions: access management to healthcare-professionals-only; text in an electronic health record is therefore richer in saliency, lower in non-specific noise and less prone to distortions. However electronic health records often have inconsistent data standardisation with structured data mixed with unstructured free text, as well as a hybrid of modern and legacy closed systems. Traditional data aggregation methods rely on gold-standard cases generated from reporting mechanisms like structured case report forms for local or national registries.

We describe an approach using real-time aggregation of keywords and key phrases from free text in electronic health records to produce a real-time signal during the Covid pandemic. This open-source system takes text from structured and unstructured fields in near-real-time from clinician-generated documentation in the electronic health records and does not require health data to be standardised into any ontology. This unstructured real-time aggregation could provide earlier warning as it avoids laboratory-sample processing delays and reduces lab undersampling bias (significant confounders during the early pandemic period).

## Results & Discussion

A query was defined producing an aggregated count of patient documents containing symptom keywords and phrases for symptomatic Covid: (“dry cough”, “pyrexia”, “fever”, “dyspnoea”, “anosmia”, “pneumonia”, “LRTI”, “lung consolidation”, “pleuritic pain” and associated synonyms, acronyms and poecilonyms) normalised against patient documents not containing these phrases or containing negations of the phrases (e.g. “no dry cough”, “no anosmia”). This was used to generate an index of signal enriched for symptom clusters of symptomatic Covid which compares favourably to the gold-standard data of laboratory samples at King’s College Hospital (KCH) and Guys and St Thomas’ Hospital (GSTT) (Figure 1). Cross-correlation peaked at 2 days before at KCH (cross-correlation r= 0.703 for 0 days lag, r=0.714 for 1 day lag, r=0.730 for 2 lag lag, r=0.725 for 3 day lag) and GSTT at 0 days before (cross-correlation r=0.234 for 0 days before, r=0.225 for 1 day lag, r=0.209 for 2 day lag) (Supplementary Figure 1).

**FIGURE 1:**
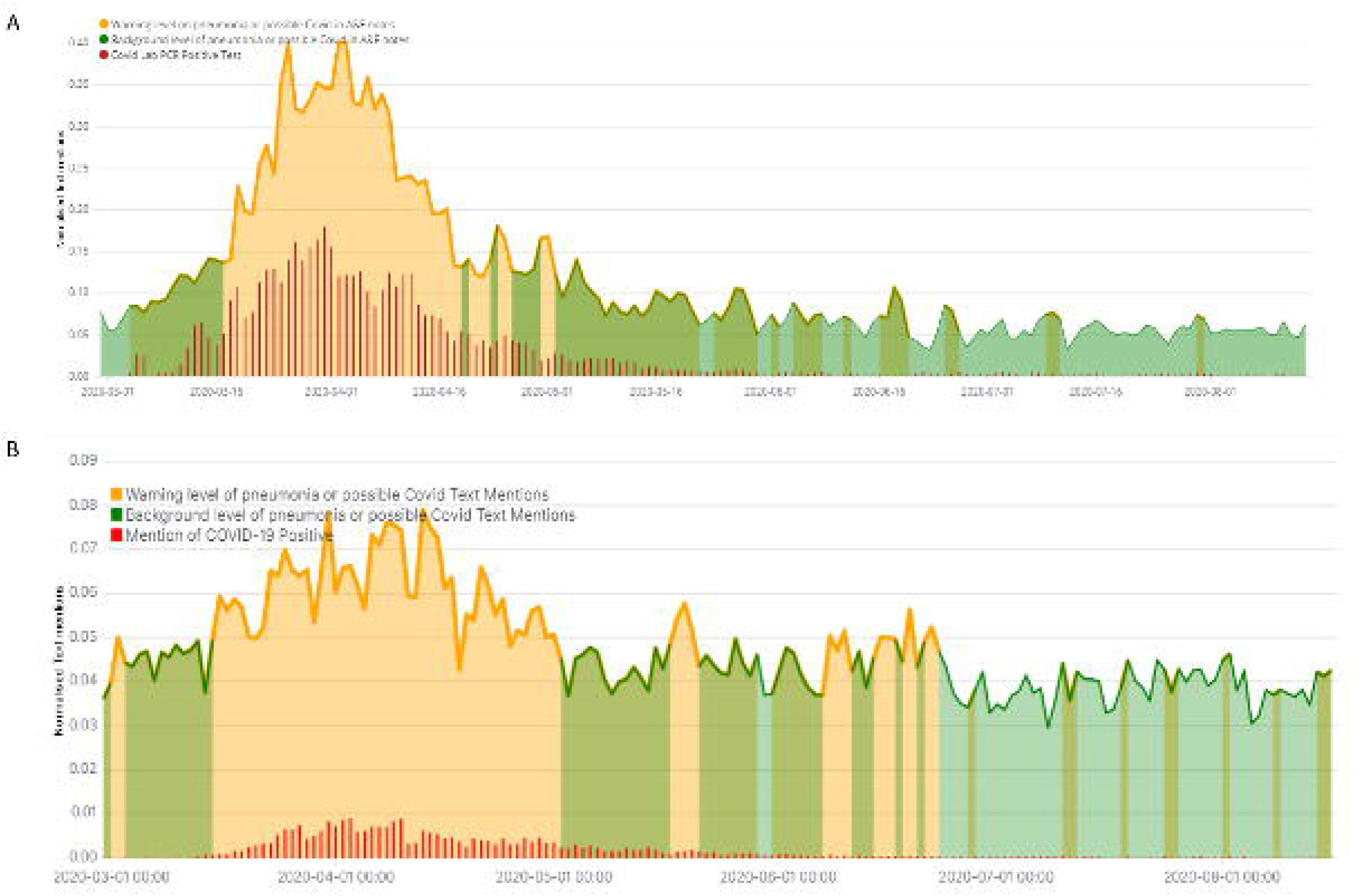
(A) Kings College Hospital (KCH) Viral Pneumonia symptom aggregator depicts the mentions of Pneumonia, Cough, and other clinical terms from the unstructured text from 28th February 2020 till 15 August 2020 with the green and orange shaded line depicting the symptom aggregator index and the red depicting the proportion of laboratory tests which are positive daily; (B) Guys & St Thomas’ Hospital (GSTT) Viral Pneumonia symptom aggregator during the same period customised to local document formats with 2 day moving averages (to reduce cyclical weekend effect). The green-orange colour scheme is used to illustrate periods of elevated incidence of the terms above a locally-defined moving-average threshold

The freetext signal is able to anticipate the awareness of Covid cases by about 1-3 days in early March 2020. As the initial surge of positive cases receded, the aggregator also declined to seasonal background levels and the persistent elevation of the index above a threshold would anticipate a further outbreak of Covid-like cases. Of note, seasonal influenza epidemics over the winters from 2018-20 were detectable due to many overlapping features (Supplementary Figure 2) indicating broad utility for viral pneumonias.

Previous attempts at text-based epidemic forecasting have been largely focused on search engine or social media trends from the public internet to generate the signal for forecasting influenza, and to measure such trends against structured public health databases at national level ^5–9^. Other approaches for forecasting have used structured manual submissions to national public health systems like the Centre for Disease Control’s Influenza-Like Illness Reports ^8^. Our approach does not attempt to forecast but acts as a noisy real-time barometer of local clinical data which is adequate for local operational use at low cost.

While our approach operates at the scale of a single healthcare organisation, it can be scaled to a whole system level either by combining locally-generated customised real-time signals from multiple organisations or by aggregating the clinical data first before generating the real-time signal. We believe the former approach is superior as a single organisation is able to customise the locally-derived signal for local operational purposes while simultaneously a wider health economy can derive utility from combining signals for other central planning purposes.

## Limitations

Ecological real-time freetext data is uncurated, and could be susceptible to distortions as certain phrases in freetext could produce artificial distortions (e.g. if ward names, job titles or operational pathways are named “Covid”). This risk is minimised in this study as the word “Covid” was not included from the signal in this study, but this could be tailored to the local context or dialect.

One can only aggregate and harness knowledge that clinicians and patients consider of sufficient relevance or saliency to record; loss of smell and anosmia has been described in Covid^10^ but this knowledge arrived formally in the scientific press on 17/05/2020. Our text aggregator was able to capture the initial signal in March 2020 (Supplementary Figure 3) and this subsequent scientific publication caused a second surge in the incidence of these phrases likely due to increasing clinician awareness.

This media-sensitive signal highlights that incident clinical language could become self-fulfilling - heightened awareness may increase the use of such words even in clinical text either through speculative differentials or checklists. We managed this by eliminating negation terms, combining multiple symptom phrases and focusing on formal documentation only (e.g. emergency department episode summaries). Nonetheless an organisation should exercise caution against disseminating the exact words or phrases used to generate the signal to minimise ‘hashtag meme’-like phenomenon amongst clinicians.

In summary, we report a natural language approach of real-time clinical data that is flexible and scalable to feed dashboards of activity for capacity planning and can be generalised across organisations to provide early warning of future pandemic surges. This low-cost approach is open-source, is not fixed into any specific electronic health record system and can be deployed at multiple organisational scales.

## Methods

### Platform

The text feed was generated by a locally-deployed Cogstack instance ^11^ at Kings’ College Hospital (KCH) NHS Foundation Trust, UK consisting of an open-source toolkit of real-time free text extracted from document stores in the electronic health record into an Elastic index (https://github.com/CogStack). A subsequent instance was deployed at Guys & St Thomas’ Hospital (GSTT) NHS Foundation Trust, UK during the Covid pandemic which shows generalisability and further customisation is ongoing to incorporate more data sources for signal optimisation.

### Text feed

A query was defined producing an aggregated count of patient documents containing symptom keywords and phrases for symptomatic Covid: (“dry cough”, “pyrexia”, “fever”, “dyspnoea”, “anosmia”, “pneumonia”, “LRTI”, “lung consolidation”, “pleuritic pain” and associated synonyms, acronyms and poecilonyms) normalised against patient documents not containing these phrases or containing negations of these phrases (e.g. “no dry cough”). This produces an index of signal which is enriched for symptom clusters of symptomatic Covid. Of note, definitive diagnostic phrases (e.g. “confirmed Covid”) were excluded from this query although these are used in practice to boost the signal.

Presence of positive COVID PCR laboratory samples was used as the gold standard signal of cases. A similar query was constructed within a parallel Cogstack instance at GSTT.

## Data Availability

The infrastructure and the authors received funding from National Institute for Health Research (NIHR) Biomedical Research Centre at South London and Maudsley NHS Foundation Trust Health Data Research UK The UK Research and Innovation London Medical Imaging and Artificial Intelligence Centre for Value Based Healthcare (AI4VBH) Innovate UK, the National Institute for Health Research (NIHR) Applied Research Collaboration South London (NIHR ARC South London) the National Institute for Health Research University College London Hospitals Biomedical Research Centre and Kings College London.

## Data Availability

To preserve privacy, aggregated counts (but no patient-level data) will be available. Any request for this will need to be reviewed first by the hospital information governance committee which includes the Caldicott Guardian and Data Protection Officer. Source code for the Cogstack platform is available at (https://github.com/CogStack). This project was conducted under public health requirements and operational service delivery of the hospital.

## Competing Statement

The infrastructure and the authors received funding from: National Institute for Health Research (NIHR) Biomedical Research Centre at South London and Maudsley NHS Foundation Trust, Health Data Research UK, The UK Research and Innovation London Medical Imaging & Artificial Intelligence Centre for Value Based Healthcare (AI4VBH), Innovate UK, the National Institute for Health Research (NIHR) Applied Research Collaboration South London (NIHR ARC South London), the National Institute for Health Research University College London Hospitals Biomedical Research Centre and King’s College London.

## Author Contribution

JT, RD, WB conceived the study design

VD, PD, VO, JT, RD performed data processing and architecture development

JT, VD, RD wrote the manuscript

VD, PD, VO, JT, RB, CB, RD performed critical review

---

**Supplementary Figure 1:**
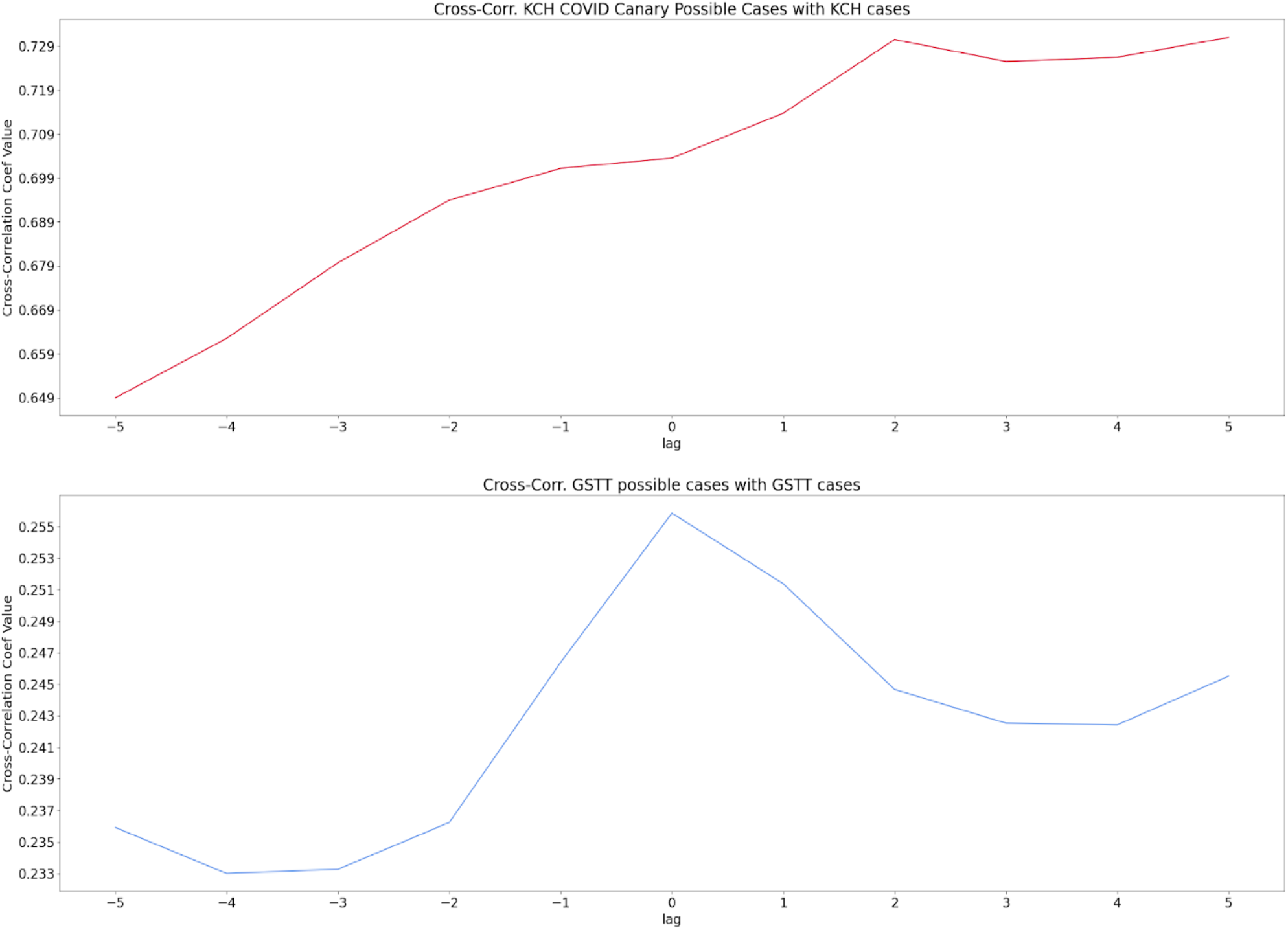
Cross-correlation of the index of the Covid signal with laboratory sample signal at KCH peaking at -2 day lag and 0 day lag

**Supplementary Figure 2:**
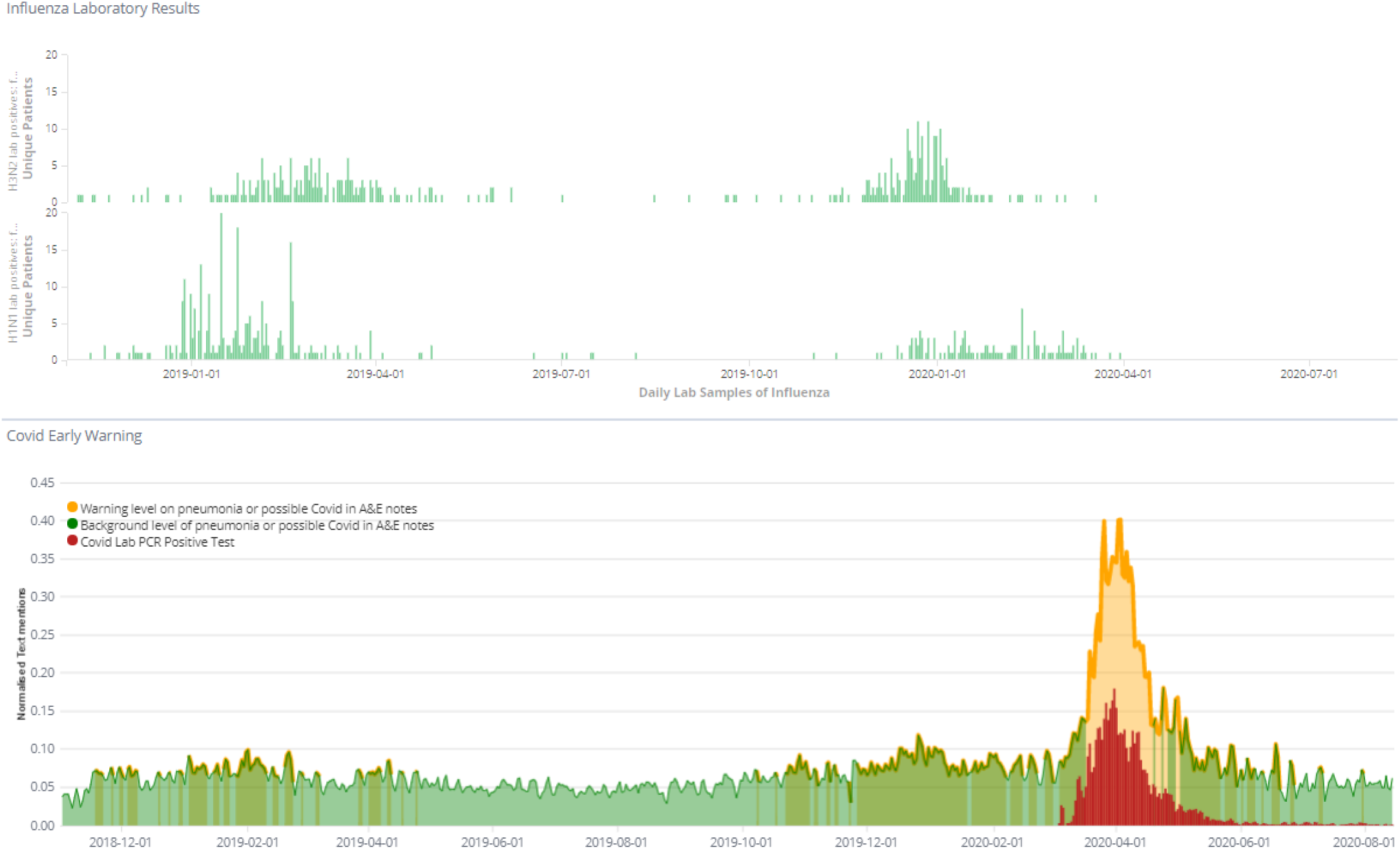
Real-time aggregation of clinical text feeds showing that the seasonal influenza symptoms are also being detected by viral pneumonia symptom aggregators (bottom) coinciding with influenza laboratory testing (top); cross-correlation for 01/01/2017 till 29/02/2020, r = 0.414 for 0 day lag, r = 0.420 for -1 day lag, r=0.348 for -2 day lag).

**Supplementary Figure 3:**
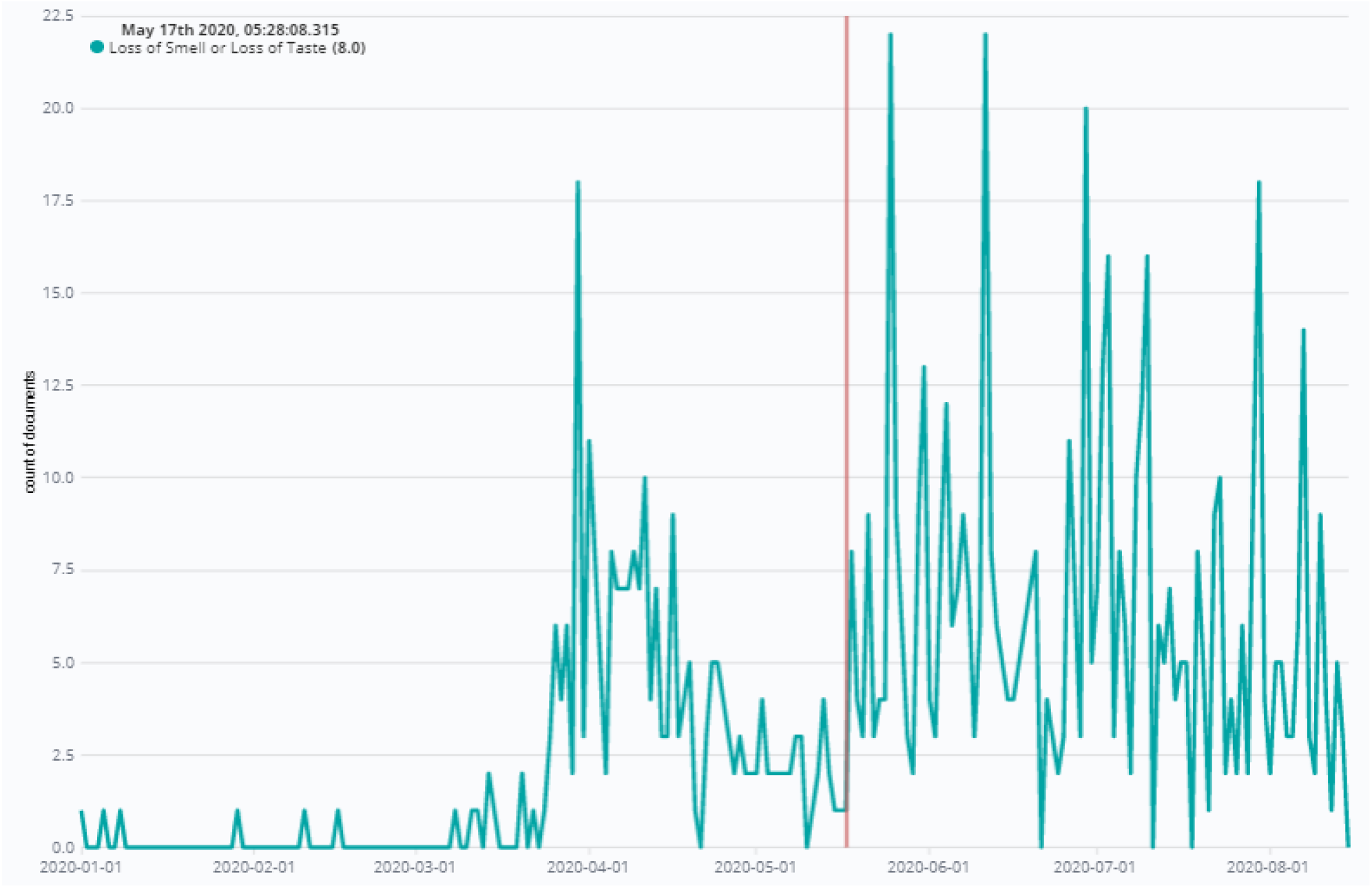
Real-time aggregation of key phrases “Anosmia”, “Loss of Taste” and “Loss of Smell” with simple negations only (e.g. “No anosmia”). Note that the detection of these terms started in March 2020 during the first Covid surge consistent with the proportion of cases, followed by a second upswell from 17/05/2020 (vertical red line) coinciding with the publication of Nature Medicine article confirming association through an app-based study ^10^.

